# A machine learning model for predicting out-of-hospital cardiac arrest incidence using meteorological, chronological, and geographical data from the United States

**DOI:** 10.1101/2023.05.08.23289698

**Authors:** Takahiro Nakashima, Soshiro Ogata, Eri Kiyoshige, Mohammad Z Al-Hamdan, Yifan Wang, Teruo Noguchi, Theresa A Shields, Rabab Al-Araji, Bryan McNally, Kunihiro Nishimura, Robert W Neumar

## Abstract

**Background:** Despite advances in pre- and post-resuscitation care, percentage of survival to hospital discharge after out-of-hospital cardiac arrest (OHCA) was extremely low. Development of an accurate system to predict the daily incidence of OHCA might provide a significant public health benefit. Here, we developed and validated a machine learning (ML) predictive model for daily OHCA incidence using high-resolution meteorological, chronological, and geographical data.

**Methods:** We analyzed a dataset from the United States that combined an OHCA nationwide registry, high-resolution meteorological data, chronological data, and geographical data. We developed a model to predict daily OHCA incidence with a training dataset for 2013–2017 using the eXtreme Gradient Boosting algorithm. A dataset for 2018–2019 was used to test the predictive model. The main outcome was the predictive accuracy for the number of daily OHCA events, based on root mean squared error (RMSE), mean absolute error (MAE), and mean absolute percentage error (MAPE). In general, a model with MAPE less than 10% is considered highly accurate.

**Results:** Among the 446,830 OHCAs of non-traumatic cause where resuscitative efforts were initiated by a 911 responder, 264,916 in the training dataset and 181,914 in the testing dataset were included in the analysis. The ML model with combined meteorological, chronological, and geographical data had high predictive accuracy in relation to nationwide incidence rate per 100,000 at the nationwide level) in the training dataset (RMSE, 0.016; MAE, 0.013; and MAPE, 7.61%) and in the testing dataset (RMSE, 0.018; MAE, 0.014; and MAPE, 6.52%).

**Conclusions:** A ML predictive model using comprehensive daily meteorological, chronological, and geographical data allows for highly precise estimates of OHCA incidence in the United States.

**Clinical Perspective:** **What is new?**

A machine learning predictive model developed with a high-resolution meteorological dataset and chronological and geographical variables predicted the daily incidence of out-of-hospital cardiac arrest (OHCA) in the U.S. population with high precision. The predictive accuracy at the state level was greater in medium and high-temperature areas than in the low-temperature area.

**What are the clinical implications?**

This predictive model revealed complex associations between meteorological, chronological, and geographic variables in relation to predicting daily incidence of OHCA. It might be useful for public health strategies in temperate regions, for example, by providing a warning system for citizens and emergency medical services agencies on high-risk days.

---

The estimated global incidence of out-of-hospital cardiac arrest (OHCA) treated by emergency medical service (EMS) is 62.0 per 100,000 person-years, with a specific incidence of 53.1 per 100,000 person-years in North America.^1^ Despite advances in pre- and post-resuscitation care, a recent systematic review showed that survival to hospital discharge after OHCA was extremely low, at 8.8% (95% confidence interval [CI], 8.2– 9.4%).^2^ Prevention and early reaction in the prehospital setting are important for improving the prognosis of patients with OHCA. Development of an accurate system to predict the daily incidence of OHCA might provide a significant public health benefit.

Several studies have shown associations between ambient temperature and cardiovascular events^3–6^ and between day of week or season and cardiovascular events.^7–12^ However, many of those studies used conventional linear regression, which might not be suitable for assessing the influence of ambient temperature on OHCA incidence and handling large amounts of high-resolution meteorological data. To date, meteorological and chronological data have not been applied to real-world practice. Machine learning (ML) can use advanced analytics to integrate multiple quantitative variables and identify associations not identified with conventional one-dimensional statistical approaches, which might allow us to develop a predictive model. Recently, our team developed a ML predictive model for daily OHCA incidence based on combined meteorological and chronological data with high accuracy for the Japanese population.^13^ In this study, we develop and validated the ML predictive model for robust estimation of daily OHCA incidence of cardiac origin for the U.S. population; the United States is close in latitude to Japan. This model was evaluated using the OHCA dataset from the Cardiac Arrest Registry to Enhance Survival (CARES) from the United States,^14^ as well as comprehensive meteorological data from National Aeronautics and Space Administration (NASA), chronological data, and geographic data.

## Methods

The study was approved by the University of Michigan Hospital’s institutional review board (HUM00189913). The requirement for written informed consent was waived because the researchers only analyzed deidentified (anonymized) data.

### Data source for out-of-hospital cardiac arrest data

OHCA data was provided by CARES, which is a prospective multicenter registry of patients with OHCA from 30 state-based registries, the District of Columbia, and more than 45 community sites in 16 additional U.S. states. It has a catchment area of approximately 175 million residents in 2021 (**Figure 1**). The design of the registry, which was established by the U.S. Centers for Disease Control and Prevention and Emory University, has been previously described.^14^ Patient-level data were collected by EMS agencies using standardized international Utstein definitions for clinical variables and outcomes to ensure uniformity.

**Figure 1.**
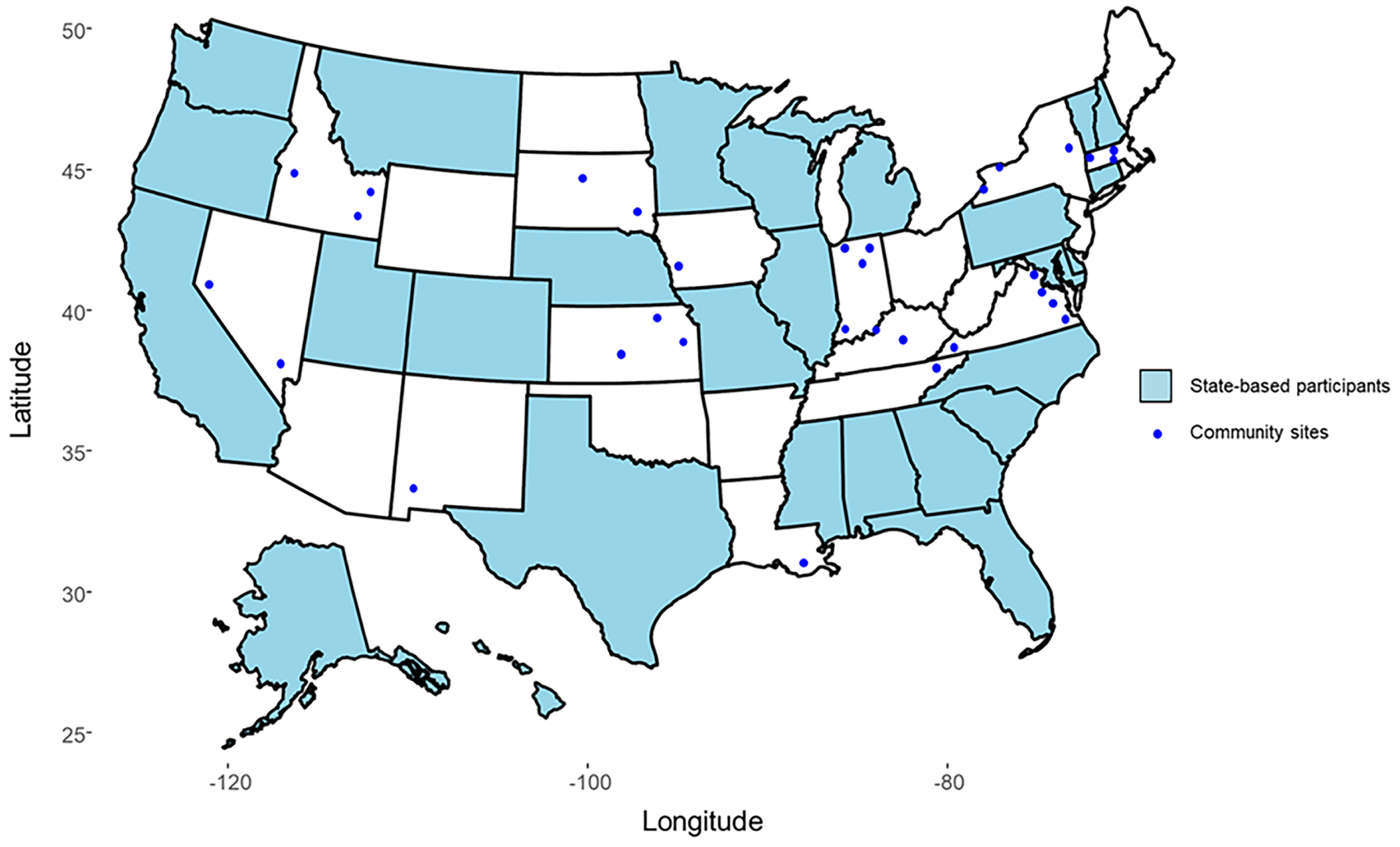
States and communities participating in CARES. CARES denotes Cardiac Arrest Registry to Enhance Survival.

CARES includes non-traumatic OHCAs where resuscitative efforts were initiated by a 911 responder. The following patient information was collected and analyzed in this study: age, sex, etiology of arrest (i.e., presumed cardiac etiology, respiratory/asphyxia, drowning/submersion, electrocution, exsanguination/hemorrhage, drug overdose, or others), and location of cardiac arrest. Each patient in the CARES registry was geocoded to a U.S. county based on the ZIP code for the location of the OHCA through a crosswalk file from the U.S. Department of Housing and Urban Development. Data were submitted in two ways: via a data entry form on the CARES website (https://mycares.net/) or daily uploads from an EMS agency’s electronic patient care record system. The CARES analyst (R.A.-A.) reviewed records for completeness and accuracy. Details of the registry are described in the **Supplemental Materials**.

### Meteorological data

We analyzed meteorological data from NASA’s North American Land Data Assimilation System, which provides hourly gridded data with 12-km spatial resolution.^15, 16^ Meteorological variables included average daily values of ambient temperature (°C), precipitation (mm), relative humidity (%), and wind speed (m/s) during the study period. Those values were averaged by EMS agency areas.

### Chronological and geographical data

Chronological variables included year (2013 was considered year 0), month (February to December as categorical variables, with January as the referent), day of the week (with Sunday as the referent), holidays, start and end of daylight saving time, and holiday season from December 24 to January 1 (categorical variable with a value of 0 or 1).

Geographical data were collected at the census tract level included median age (four categories: <35, 35–39, 40–44, and ≥45 year, near its quartiles), proportion of men (two categories: <50% or ≥50%, which approximate the median), race (proportion of Blacks [three categories: <1%, 1–6%, and ≥6%, near its tertiles], proportion of Asians [three categories: <1%, 1–2%, and ≥3%, near its tertiles]), proportion of individuals with a high school diploma or higher (three categories: <55%, 55–60%, 60–64%, and ≥65%, near its quartiles), unemployment rate (four categories: <4%, 4–4.9%, 5–6%, and ≥6%, near its quartiles), percentage of individuals living below the poverty level (four categories: <7%, 7–10%, 11–14%, and ≥15%, near its quartiles), average household size (four categories: <2.4, 2.4–2.55, 2.56–2.69, and ≥2.7, near its quartiles), and population per square mile (four categories: <500, 500–1,699, 1700–3,200, and 3,200 and over, near its quartiles). The geographical data at the census tract level were merged into EMS agency areas. In cases where an EMS agency area was covered by multiple census tract areas, we used the geographical from the tract with the highest number of OHCA cases among the several tract areas.

### Data management and development of predictive models

We matched the CARES data, meteorological data, chronological data, and geographic data between January 1, 2013, and December 31, 2019 at the hourly level based on the time of the emergency call. We classified data from January 1, 2013 to December 31, 2017 in this merged dataset as the training dataset for developing the predictive model. Of EMS agencies in the training dataset, 30% were used as the validation dataset for selecting hyperparameters. Data from January 1, 2018 to December 31, 2019 were used as the testing dataset for assessing whether the predictive model can work in other years.

To develop predictive model for the daily incidence of OHCA, we used the eXtreme Gradient Boosting (XGBoost) gradient boosting algorithm.^17, 18^ We selected XGBoost hyperparameters that maximized prediction performance in the validation dataset. In other words, we minimized root mean squared error (RMSE) by developing a model in the dataset between 2013 and 2017 in 70% of the participating agency areas and checking its prediction performances according to its hyperparameters in the validation dataset (i.e., data between 2013 and 2017 in the remaining 30% of agency areas). Population size for each agency area was included in the XGBoost algorithm as an offset term. We assessed prediction performance of the model developed with the testing dataset (i.e., data between 2018 and 2019 from all participating agency areas).

### Primary and secondary outcomes

The primary outcome was predictive accuracy of OHCA incidence rate per 100,000 at nationwide level of the predictive model based on RMSE, mean absolute error (MAE), and mean absolute percentage error (MAPE), which are generally used as measures of predictive accuracy for a forecasting method. The secondary outcome was predictive accuracy of OHCA incidence rate per 100,000 at the state level, which was limited to 24 state-based registries.

### Statistical analysis

We performed a third-step analysis. First, we examined the concordance between the predicted incidence of OHCA based on the ML model and the observed incidence of OHCA in the testing dataset. Next, we investigated important predictors for predicting the OHCA incidence in the developed prediction model. Finally, we assessed the predictive accuracy of the ML model stratified into low-, intermediate-, and high-temperature areas, further divided into summer (June–August) and winter (December– February). Low-, intermediate-, and high-temperature areas were defined as regions with mean ambient temperature in the 25th percentile or lower, in the 25–75th percentiles, and 75th percentile or higher, respectively.

The characteristics of the present dataset were summarized with medians and interquartile ranges (IQRs) for continuous variables, and numbers and percentages for categorical variables by area and day in the training and testing datasets. We evaluated the predictive accuracy of the predictive models based on RMSE, MAE, and MAPE between predicted values calculated with the predictive models and observed daily OHCA incidence at the EMS agency level. RMSE and MAE reflect the average magnitude of differences between predicted values and observed values. RMSE and MAE can range from zero to infinity. Lower RMSE and MAE values indicate higher predictive performance. MAPE is an average of the absolute values of errors divided by observed values. MAPE ranges from zero to infinity. Lower MAPE values indicate higher model predictive performance. In general, MAPE less than 10% is considered highly accurate predicting.^19^ Formulas are as follows;

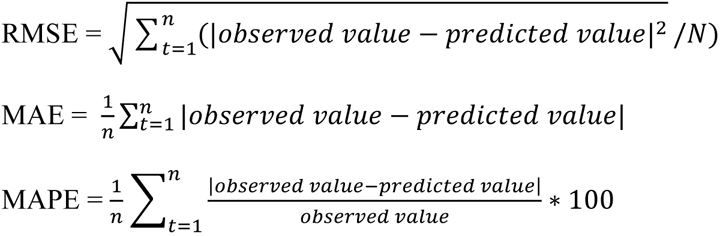

To show important predictors of the OHCA incidence in the developed prediction model, we used the Shapley Additive Explanations (SHAP) values summarizing contribution of each predictor to the predicted value of an instance.^20^ For a given set of feature values, a SHAP value reflects how much a single variable, in the context of its interaction with other variables, contributes to the difference between the actual prediction and the mean prediction.

All statistical analyses were performed with R statistical software, version 4.1.2 (https://www.R-project.org/) and the xgboost package for R, version 1.5.0.1 (https://CRAN.R-project.org/package=xgboost). Missing values for continuous and categorical variables were, respectively, imputed by a median value for each continuous variable and treated as a missing category. These missing procedures work well when using XGBoost due to the nature of decision tree algorithms.

## Results

### Characteristics of the training and testing datasets

From the CARES registry, 446,830 EMS-treated OHCAs of non-traumatic cause between 2013 and 2019 were matched with meteorological data; there were 264,916 cases in the training dataset. There were 181,914 cases in the testing dataset. The characteristics of the datasets are summarized in **Table 1**. Between 2013 and 2019, the median annual incidence of OHCA increased from 58.6 to 76.3 per 100,000 person-years. The median age of OHCA onset increased from 64 (IQR, 52–77) to 65 years (IQR, 53–76). The proportion of males increased from 61% to 62%.

**Table 1.** Characteristics of daily data in the training dataset (2013–2017) and testing dataset (2018–2019)

| Variable | Training dataset<br>(n=264,916) |  | Testing dataset<br>(n=181,914) |
| --- | --- | --- | --- |
|  | 2013–2015 | 2016–2017 | 2018–2019 |
| **Demographic variables** |  |  |  |
| Number of participating EMS agencies | 575 | 845 | 1222 |
| OHCA incidence, per 100,000 person-years | 58.6 | 70.4 | 76.3 |
| **Patient characteristics** |  |  |  |
| Age, years, median (IQR) | 64 (52, 77) | 64 (52, 76) | 65 (53, 76) |
| Male sex, n (%) | 81,903 (61) | 86,761 (62) | 115,674 (62) |
| Race, n (%) |  |  |  |
| White | 61,961 (46) | 67,537 (48) | 94,518 (51) |
| Black | 27,408 (20) | 31,402 (22) | 43,049 (23) |
| Asian | 2,531 (2) | 2,903 (2) | 4,046 (2) |
| Hispanic/Latino | 7,952 (6) | 7,506 (5) | 12,199 (7) |
| Other | 34,529 (26) | 31,074 (23) | 32,212 (17) |
| First documented rhythm, n (%) |  |  |  |
| Shockable | 27,547 (20) | 26,599 (19) | 34,718 (19) |
| Non-shockable | 106,823 (79) | 113,810 (81) | 151,279 (81) |
| **Meteorological variables** |  |  |  |
| Ambient temperature, °C |  |  |  |
| Mean value within a day |  |  |  |
| Low-temperature area | 8.9 (−0.1, 18.1) | 9.6 (0.2, 18.0) | 7.9 (−0.7, 18.3) |
| Intermediate temperature area | 11.9 (4.5, 19.5) | 11.9 (4.4, 19.4) | 11.6 (3.99, 20.0) |
| High-temperature area | 17.4 (10.8, 24.0) | 17.1 (10.7, 23.3) | 17.6 (10.7, 24.5) |
| Differences between maximum and minimum values within a day |  |  |  |
| Low-temperature area | 9.4 (6.8, 12.1) | 8.8 (6.1, 11.2) | 9.6 (7.0, 12.4) |
| Intermediate temperature area | 9.2 (6.5, 11.8) | 9.0 (6.2, 11.6) | 9.9 (7.4, 12.7) |
| High-temperature area | 10.3 (7.8, 12.7) | 9.6 (7.1, 12.0) | 10.6 (7.8, 13.6) |
| Precipitation during the previous hour, mm |  |  |  |
| Low-temperature area | 0.00 (0.00, 0.06) | 0.00 (0.00, 0.09) | 0.00 (0.00, 0.09) |
| Intermediate temperature area | 0.00 (0.00, 0.07) | 0.00 (0.00, 0.09) | 0.00 (0.00, 0.10) |
| High-temperature area | 0.00 (0.0, 0.10) | 0.00 (0.00, 0.05) | 0.00 (0.00, 0.05) |
| Wind speed, m/s |  |  |  |
| Low-temperature area | 3.2 (2.1, 4.5) | 3.5 (2.3, 4.8) | 3.4 (2.2, 4.8) |
| Intermediate temperature area | 3.0 (2.0, 4.3) | 3.4 (2.3, 4.7) | 3.2 (2.2, 4.5) |
| High-temperature area | 3.1 (2.2, 4.2) | 3.2 (2.2, 4.3) | 3.1 (2.1, 4.3) |
| Relative humidity, % |  |  |  |
| Low-temperature area | 70.8 (55.5, 81.6) | 77.6 (70.2, 84.7) | 78.7 (69.9, 85.5) |
| Intermediate temperature area | 74.0 (62.2, 82.8) | 75.8 (67.7, 83.7) | 77.9 (69.8, 84.5) |
| High-temperature area | 69.3 (55.7, 79.1) | 72.3 (63.3, 81.0) | 74.1 (61.7, 82.1) |

Continuous values are presented as medians (IQR).
\*See **eFigure 1 in the Supplement** for more information.
†Snowfall values were calculated for the winter (December–February).
IQR denotes interquartile range; hPa, hectopascal; OHCA, out-of-hospital cardiac arrest.

The median of the mean ambient temperature within a day decreased from 8.9°C (IQR, −0.1 to 18.1) to 7.9°C (IQR, −0.7 to 18.3) in the low-temperature area. This trend was not observed in the intermediate or high-temperature areas. Differences between maximum and minimum ambient temperatures within a day (diurnal temperature range) increased from 9.4°C (6.8–12.1) to 9.6°C (7.0–12.4) in the low-temperature area, from 9.2°C (6.5–11.8) to 9.9°C (7.4–12.7) in the intermediate-temperature area, and from 10.3°C (7.8–12.7) to 10.6°C (7.8–3.6) in the high-temperature area. Relative humidity also increased throughout the study period in all areas. The incidence of OHCA by meteorological condition is shown in **Figure 2**.

**Figure 2.**
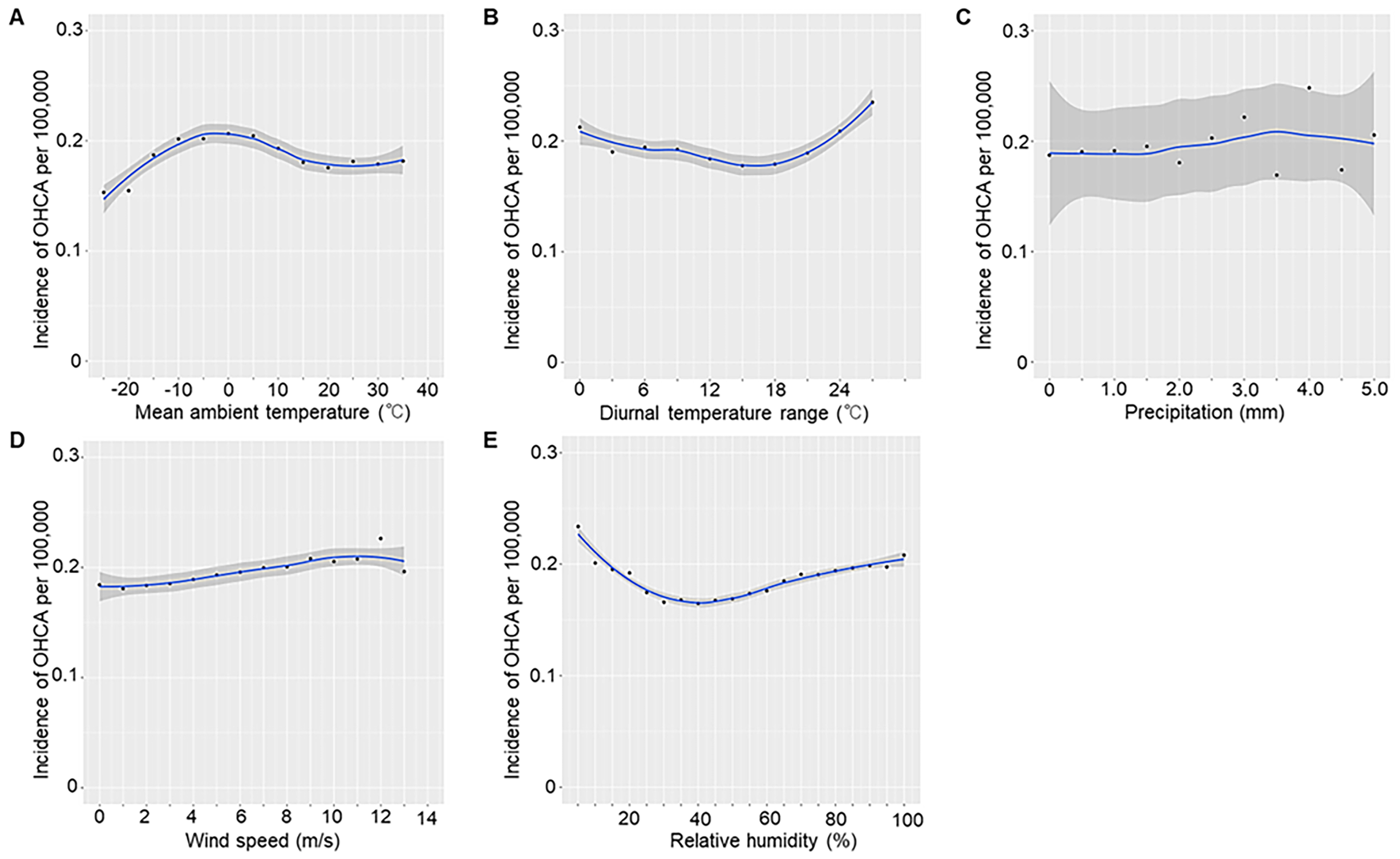
Incidence of out-of-hospital cardiac arrest by meteorological condition. The plots indicate the mean daily OHCA incidence per 100,000 persons. OHCA denotes out-of-hospital cardiac arrest.

### Predictive performance of the models

Predicted and observed incidence of OHCA with a cardiac origin for each model are shown in **Figure 3**. The ML-predicted model, which incorporated meteorological, chronological, and geographical variables, was able to accurately predict daily fluctuations and a significant increase in OHCA incidence at the nationwide level, demonstrating good concordance between predicted and observed values. The error rate was less than 0.05 per 100,000 persons per day. The predictive accuracy of the models is shown in **Table 2**. At the nationwide level, the ML predictive model had high predictive accuracy in the training dataset (RMSE, 0.016; MAE, 0.013; and MAPE, 7.61%) and the testing dataset (RMSE, 0.018; MAE, 0.014; and MAPE, 6.52%). At the state level, the model had RMSE of 0.216 and MAE of 0.128 with the training dataset and RMSE of 0.209 and MAE of 0.131 with the testing dataset. At the agency level, the model had RMSE of 1.018 and MAE of 0.287 with the training dataset and RMSE of 1.290 and MAE of 0.343 with the testing dataset.

**Figure 3.**
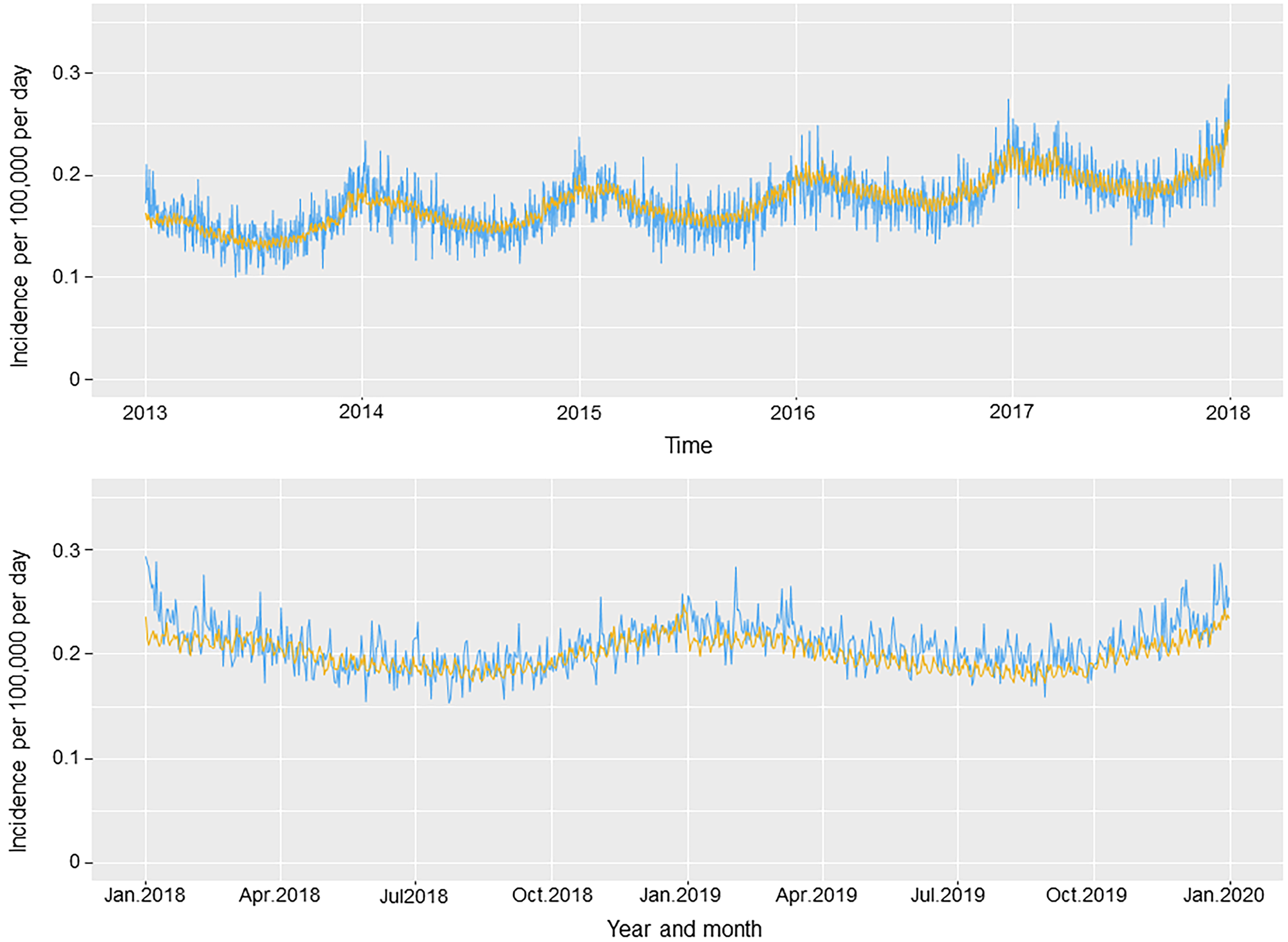
Observed versus predicted incidence of out-of-hospital cardiac arrest. The light blue lines indicate the observed total number of out-of-hospital cardiac arrests per day in the United States. The yellow lines indicate the predicted number based on combined meteorological, chronological, and geographical variables. Apr denotes April; Jan, January; Jul, July; Oct, October.

**Table 2.** Performance of the predictive model for out-of-hospital cardiac arrest based on combined meteorological and chronological data.

| Measure of predictive model performance | Training dataset | Testing dataset |
| --- | --- | --- |
| Primary outcomes |  |  |
| Nationwide level |  |  |
| RMSE, incidence rate per 100,000 per day | 0.016 | 0.018 |
| MAE by day, incidence rate per 100,000 per day | 0.013 | 0.014 |
| MAPE by day, %<sup>a</sup> | 7.61 | 6.52 |
| Secondary outcomes |  |  |
| State level<sup>b</sup> |  |  |
| RMSE, incidence rate per 100,000 per day | 0.216 | 0.209 |
| MAE by day, incidence rate per 100,000 per day | 0.128 | 0.131 |
| MAPE by day, %<sup>a</sup> | NA | NA |
| Agency level |  |  |
| RMSE, incidence rate per 100,000 per day | 1.018 | 1.290 |
| MAE by day, incidence rate per 100,000 per day | 0.287 | 0.343 |
| MAPE by day, %<sup>a</sup> | NA | NA |

- 419 a. In general, MAPE <10% is considered highly accurate predicting; 10–20%, good predicting; 20–50%, reasonable predicting; and >50%, inaccurate predicting.<sup>22</sup> MAPE could not be calculated at the state or agency level because there were days with no cardiac arrests. - 422 b. State-level calculations were limited to 24 states that participated in state-based data collection through 2017 (training dataset). - 424 MAE denotes mean absolute error; MAPE, mean absolute percentage error; ML machine learning; NA, not applicable; RMSE, root mean squared error.

### Contribution of each predictor to the predicted value of OHCA incidence

Importance of meteorological, chronological, and geographical variables in the ML predictive model is shown in **Figure 4**. In general, geographical and meteorological variables contributed more to the ML model than chronological variables. With regards to meteorological variables, lower mean ambient temperature within a day was the variable most strongly contributing to the predicted OHCA incidence, followed by mean relative humidity and larger diurnal temperature range. Among chronological variables, year, day of week, and holiday contributed more strongly to the predicted OHCA incidence than month, but their contributions were relatively small. Among geographic variables, percentage of Black persons, percentage of individuals living under the poverty level, unemployment rate, median age, and population per square mile contributed strongly to the predicted OHCA incidence.

**Figure 4.**
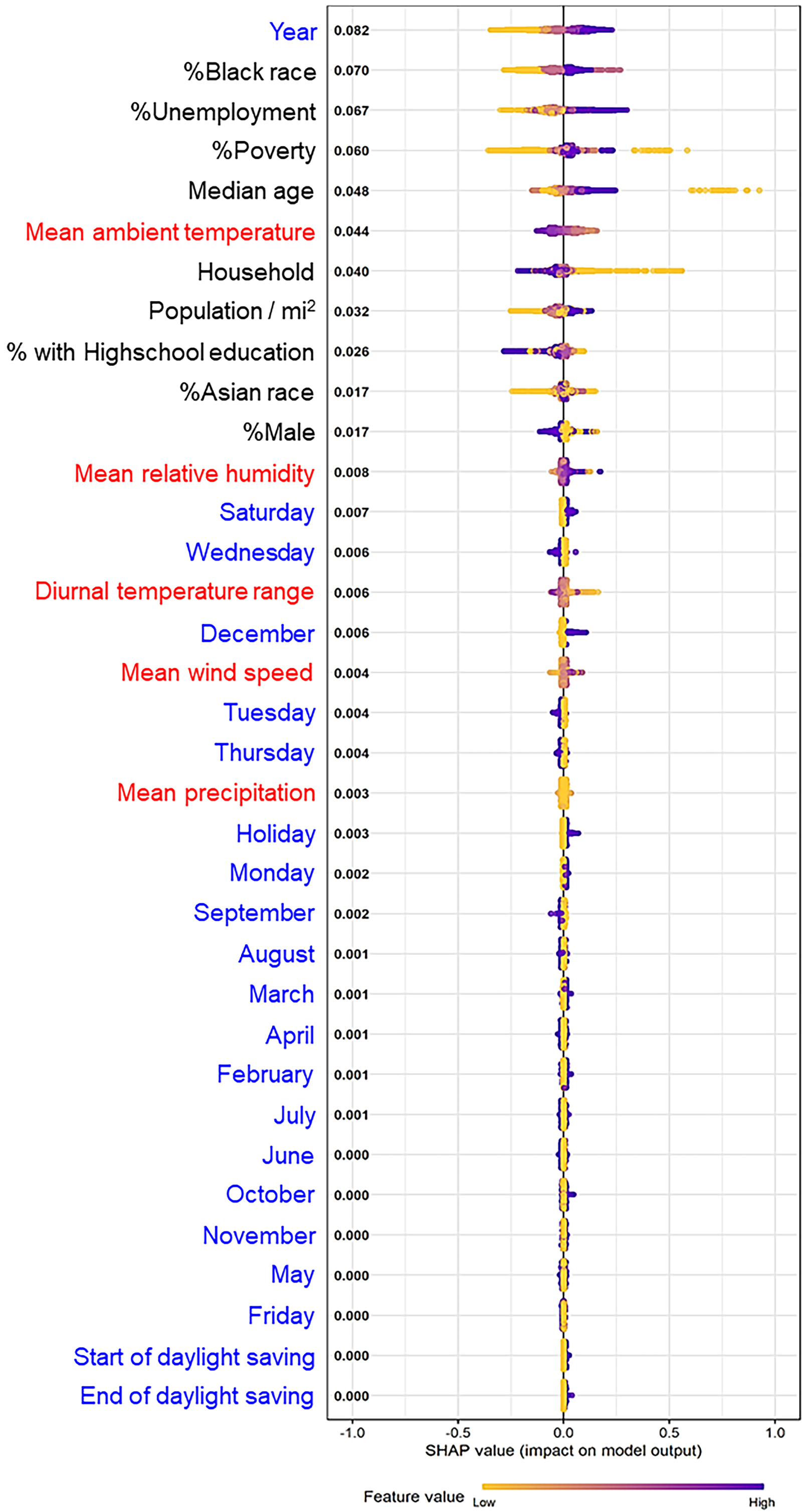
Importance of meteorological, chronological, and geographical variables in a machine learning predictive model. This figure shows a variable importance plot for meteorological variables (red), chronological variables (blue), and geographical variables (black) in a machine learning predictive model using XGBoost. The yellow to purple dots in each row represent low to high values for each predictor normally scaled. The *x*-axis shows the Shapley value, indicating the variable’s impact on the model. Positive SHAP values tend to drive predictions toward an OHCA event and negative SHAP values tend to drive the prediction toward no OHCA event. * In the model, 2013 was considered year 0. OHCA denotes out-of-hospital cardiac arrest; SHAP, Shapley Additive Explanations; XGBoost, eXtreme Gradient Boosting.

### Predictive performance based on annual average of daily mean ambient temperature

We compared the predictive accuracy of the ML model stratified by annual average of ambient temperature (**Table 3**). Population and OHCA incidence per 100,000 person-years were higher in the high-temperature area, followed by the intermediate-temperature area and low-temperature area. Predictive accuracy was higher in the intermediate- and high-temperature areas than in the low-temperature area in the training and testing datasets, as was OHCA incidence. However, there was not much difference in the predictive accuracy between summer and winter in any area.

**Table 3.**
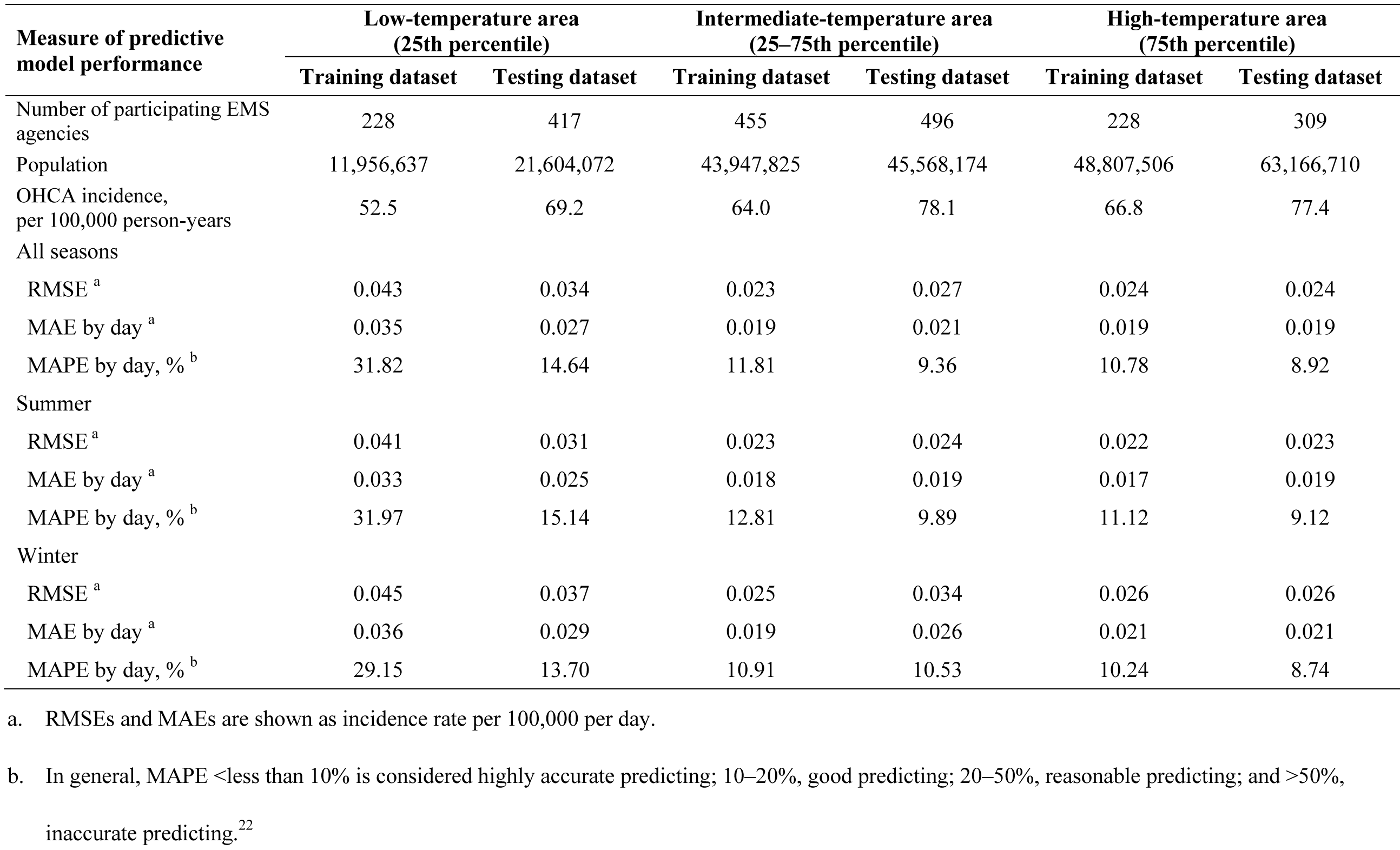
Accuracy of the predictive model for out-of-hospital cardiac arrest stratified by temperature.

| Measure of predictive model performance | Low-temperature area<br>(25th percentile) |  | Intermediate-temperature area<br>(25–75th percentile) |  | High-temperature area<br>(75th percentile) |  |
| --- | --- | --- | --- | --- | --- | --- |
|  | Training dataset | Testing dataset | Training dataset | Testing dataset | Training dataset | Testing dataset |
| Number of participating EMS agencies | 228 | 417 | 455 | 496 | 228 | 309 |
| Population | 11,956,637 | 21,604,072 | 43,947,825 | 45,568,174 | 48,807,506 | 63,166,710 |
| OHCA incidence, per 100,000 person-years | 52.5 | 69.2 | 64.0 | 78.1 | 66.8 | 77.4 |
| All seasons |  |  |  |  |  |  |
| RMSE <sup>a</sup> | 0.043 | 0.034 | 0.023 | 0.027 | 0.024 | 0.024 |
| MAE by day <sup>a</sup> | 0.035 | 0.027 | 0.019 | 0.021 | 0.019 | 0.019 |
| MAPE by day, % <sup>b</sup> | 31.82 | 14.64 | 11.81 | 9.36 | 10.78 | 8.92 |
| Summer |  |  |  |  |  |  |
| RMSE <sup>a</sup> | 0.041 | 0.031 | 0.023 | 0.024 | 0.022 | 0.023 |
| MAE by day <sup>a</sup> | 0.033 | 0.025 | 0.018 | 0.019 | 0.017 | 0.019 |
| MAPE by day, % <sup>b</sup> | 31.97 | 15.14 | 12.81 | 9.89 | 11.12 | 9.12 |
| Winter |  |  |  |  |  |  |
| RMSE <sup>a</sup> | 0.045 | 0.037 | 0.025 | 0.034 | 0.026 | 0.026 |
| MAE by day <sup>a</sup> | 0.036 | 0.029 | 0.019 | 0.026 | 0.021 | 0.021 |
| MAPE by day, % <sup>b</sup> | 29.15 | 13.70 | 10.91 | 10.53 | 10.24 | 8.74 |

a. RMSEs and MAEs are shown as incidence rate per 100,000 per day.
b. In general, MAPE <less than 10% is considered highly accurate predicting; 10–20%, good predicting; 20–50%, reasonable predicting; and >50%, inaccurate predicting.<sup>22</sup>

## Discussion

In this study, using an ML predictive model developed with the combination of meteorological, chronological, and geographic variables, we successfully predicted the daily incidence of OHCA due to a cardiac origin in the United States with high precision at the nationwide, state, and agency level, respectively. Lower mean ambient temperature within a day and larger difference between maximum and minimum ambient temperatures within a day were strongly associated with daily OHCA incidence.

An association between ambient temperature and incidence of cardiovascular events has been previously reported.^3–10^ However, since these studies focused on ambient temperature or season alone, diversity in climate (comprehensive meteorological variables), chronological variables, and geography were not considered. Recently, our team reported that an ML predictive model for OHCA incidence in Japan based on a comprehensive meteorological dataset and chronological variables had high predictive accuracy.^13^ In the present study, when geographical data were added to the predictive model, we achieved a higher predictive accuracy in the U.S. population than in the Japanese population (MAPE, 6.52% vs. 7.79%). Notably, patients with OHCA in the United States were more likely to be younger (64 vs. 80 years) or male (62% vs. 57%) than patients with OHCA in Japan. When chronological and geographic data were added to the model, the association between OHCA incidence and meteorological conditions can be more widely generalizable for the general population, even in countries with a different OHCA profile as long they are at a similar latitude range.

We found that geographic data as well as race, socioeconomic disparities, and age are strongly associated with OHCA incidence in a SHAP analysis. However, while geographic characteristics were related to differences between counties, they are not relevant to daily fluctuations in OHCA incidence because those variables do not change throughout the year. For meteorological variables, mean ambient temperature in a day and a larger diurnal temperature range were associated with the incidence of OHCA with a cardiac origin. Wolf et al., who analyzed 9,801 patients from a cardiovascular disease registry in Germany, reported that a drop in ambient temperature of 10°C or more within a day and ambient temperature itself are associated with myocardial infarction.^5^ A similar trend was observed recently in our study of a ML predictive model for OHCA in the Japanese population.^13^ The present results were also consistent with the previously described relationship between ambient temperature and OHCA incidence. Race and socioeconomic disparities might emphasize the relationship between meteorological conditions and OHCA incidence.

The predictive accuracy of the model was generally lower at the state level than at the nationwide level. Our ML predictive model had variations in predictive accuracy across states. Analyses stratified by annual average ambient temperature showed that predictive accuracy was the lowest in the low-temperature area, while the season variable (summer or winter) did not substantially change predictive accuracy. These results were partially explained by the population in the participating area. Collecting more samples improves our ML model. In addition, populations residing in the low-temperature area might be more habituated and better able to cope with climate change, such as through building insulation and lifestyle habits. Curriero et al. reported a latitude dependence of the temperature–mortality relationship in their analysis based on 11 eastern cities in the United States.^21^ More effective adaption to colder temperature was observed in cities that are further north. In order to be more practical, it needs to be further improved to predict OHCA incidence within a medical catchment area. Weather forecasts can predict meteorological conditions approximately 10 days ahead. Thus, a predictive model based on meteorological data might allow EMS agencies and hospitals to prepare and reallocate medical resources, which leads to more rapid transport and advanced post-arrest care. A future prospective study to evaluate the effectiveness of this approach is needed.

This study has several inherent limitations. First, although the CARES registry is the largest database of OHCA in the United States, it covered approximately 53% of catchment areas as of 2022. CARES only includes EMS-treated OHCA. This underestimates the overall incidence of OHCA, but is most relevant population on which to focus public health interventions. Second, our data did not address the potential variability in patients’ preexisting medical conditions. Third, the predictability of future OHCA events will depend on the accuracy of meteorological data. Finally, external testing in other developed countries was not performed.

## Conclusion

An ML predictive model using multiple meteorological, chronological, and geographical variables could predict the incidence of OHCA with a cardiac origin with high precision in the U.S. population. This predictive model might be useful for public health prevention strategies in temperate regions, which can also be applied even in countries with a different OHCA profile.

## Data Availability

The CARES registry and meteorological data were used with permission for this study. Data sharing outside of the research team was not permitted. However, the analytic code in R can be shared upon request.

https://mycares.net/

## Acknowledgments

We thank all the EMS agencies for their cooperation in establishing and maintaining the CARES registry.

## Contributors

T. Nakashima contributed to formulation of the study design, interpretation of the results, and preparation of the final report. T. Noguchi and R.W. Neumar contributed to the final report. K. Nishimura, S. Ogata, and Kiyoshige contributed to data analysis. B. McNally, R. Al-Araji, and T. Shields contributed to formulation of the concept of the CARES registry, data collection, and data management. M. Al-Hamadan and W. Yifan contributed to data collection and analysis of meteorological data. All authors approved the final version.

## Funding

This research was partially supported by a Grant-in-Aid for Young Scientists (A) (20K17914) from the Japan Society for the Promotion of Science. All authors had full access to all datasets. The corresponding author had the ultimate responsibility for the decision to submit for publication.

## Competing Interests

All authors declare no competing interests.

## Notes

### Competing Interest Statement

The authors have declared no competing interest.

